## Supplementary Figures for "Polygenic risk score for ulcerative colitis predicts immune checkpoint inhibitor-mediated colitis"

**Supplementary Figure 1: Receiver operating curves for (i) PRS$_{CD}$, and (ii) PRS$_{UC}$, respectively in UK Biobank (Testing)**

(i) **PRS$_{CD}$**

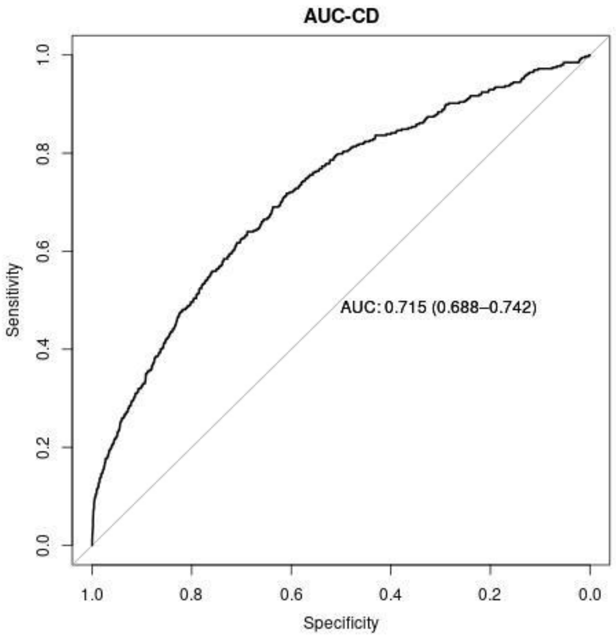


(ii) **PRS$_{UC}$**

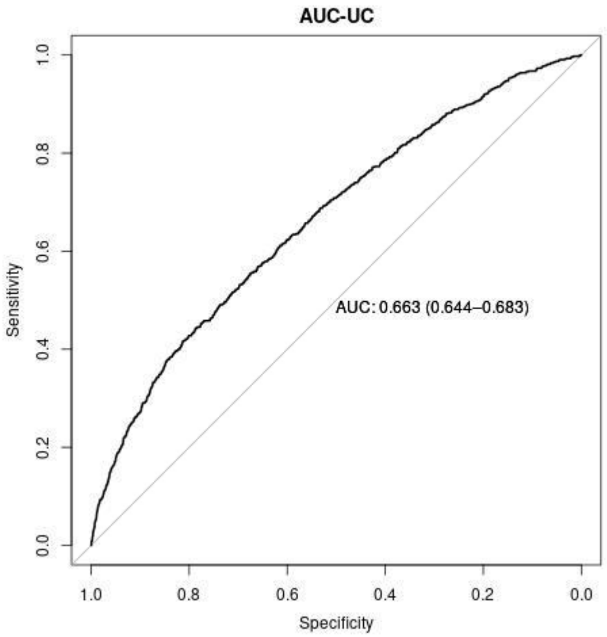

**Supplementary Figure 2: Receiver operating curves for (i) $PRS_{CD}$, and (ii) $PRS_{UC}$, respectively in BioVU (Validation cohort)**

(i) **$PRS_{CD}$**

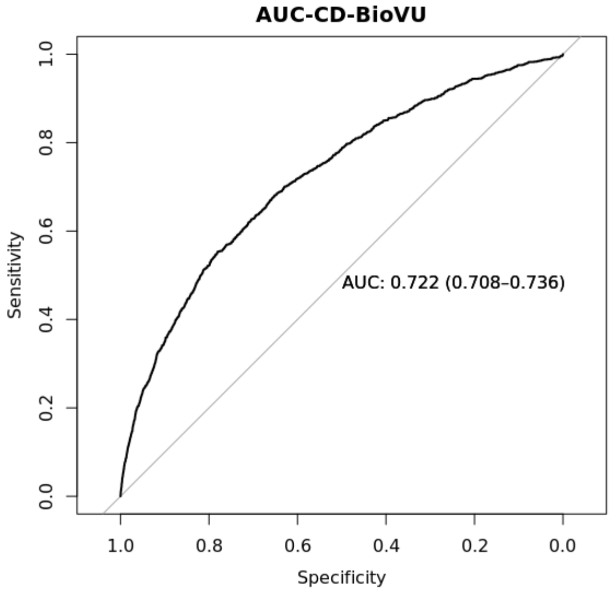


(ii) **$PRS_{UC}$**

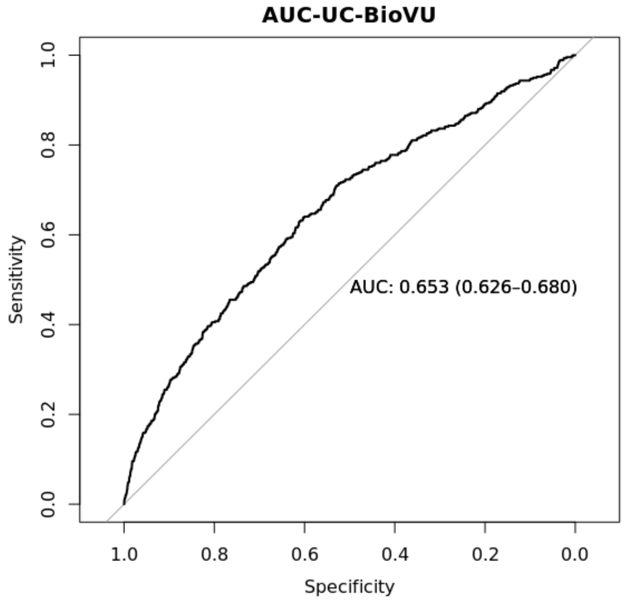

**Supplementary Figure 3: Differences in the mean $PRS_{UC}$ by immune checkpoint inhibitor-mediated colitis (All-grade and severe) in the GeRI cohort**

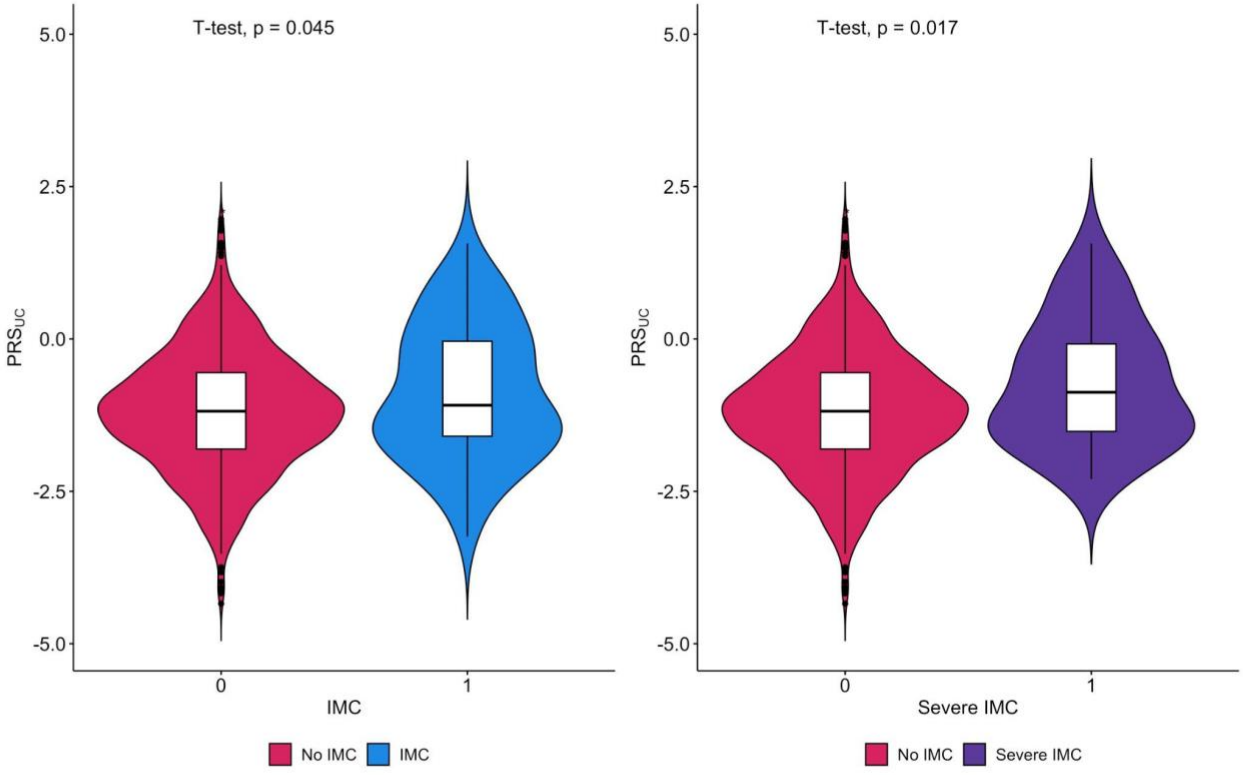

**Supplementary Figure 4: Associations of known ulcerative colitis-associated HLA markers (frequency ≥0.01) with all-grade immune checkpoint inhibitor-mediated colitis in the GeRI cohort.**

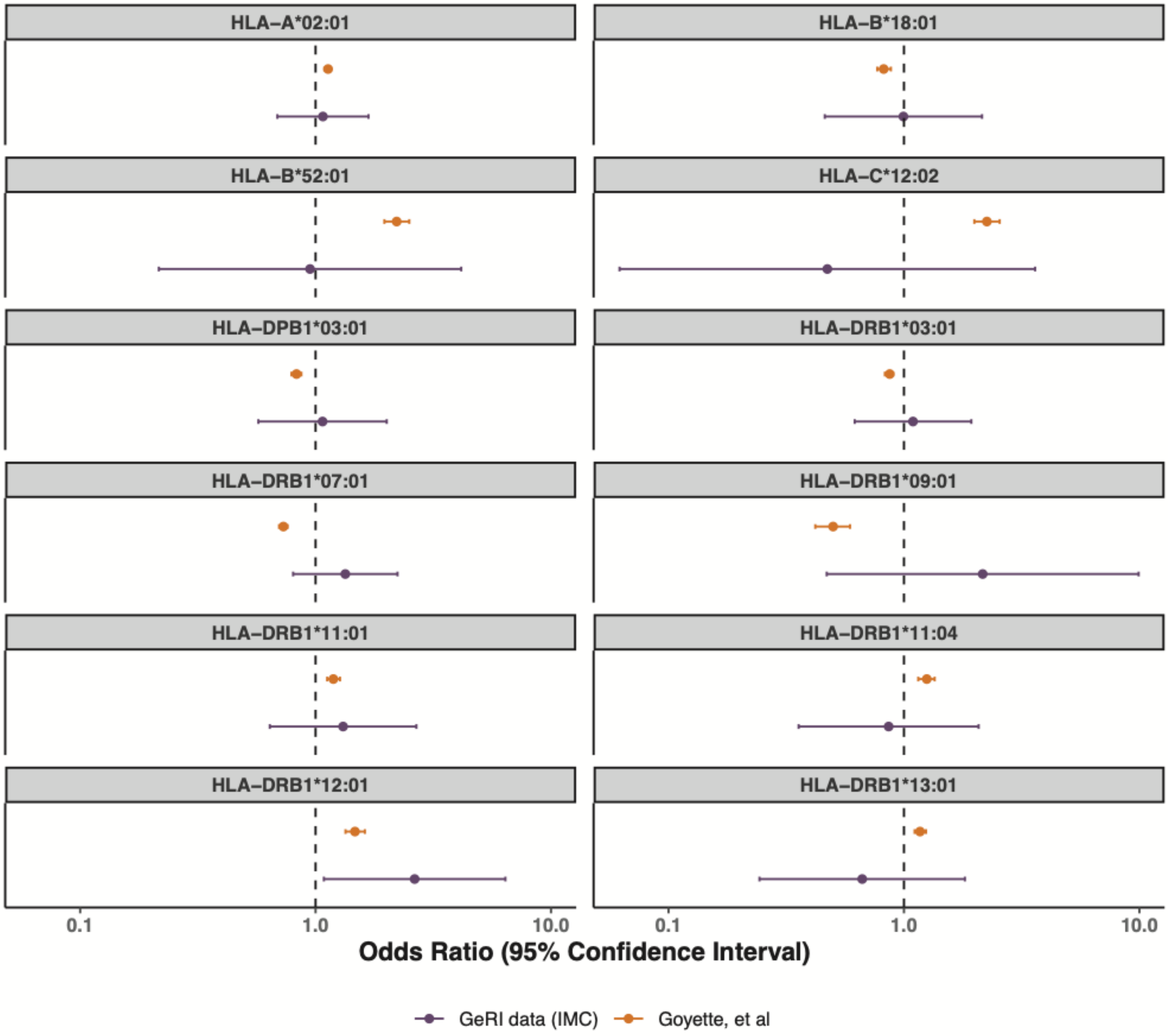

**Supplementary Figure 5: Immune checkpoint inhibitor-mediated colitis (IMC) as a predictor of progression-free survival (PFS) in the GeRI cohort (i) All-grade IMC, (iii) Severe IMC. Kaplan–Meier survival curves are unadjusted and compare those who had an IMC (all-grade or severe) with those who did not have an IMC (No IMC). The p-values in the graph represent the log-rank p-values and the dotted line represents median survival time. Graphs are cox proportional hazards models with 90-day landmark.**

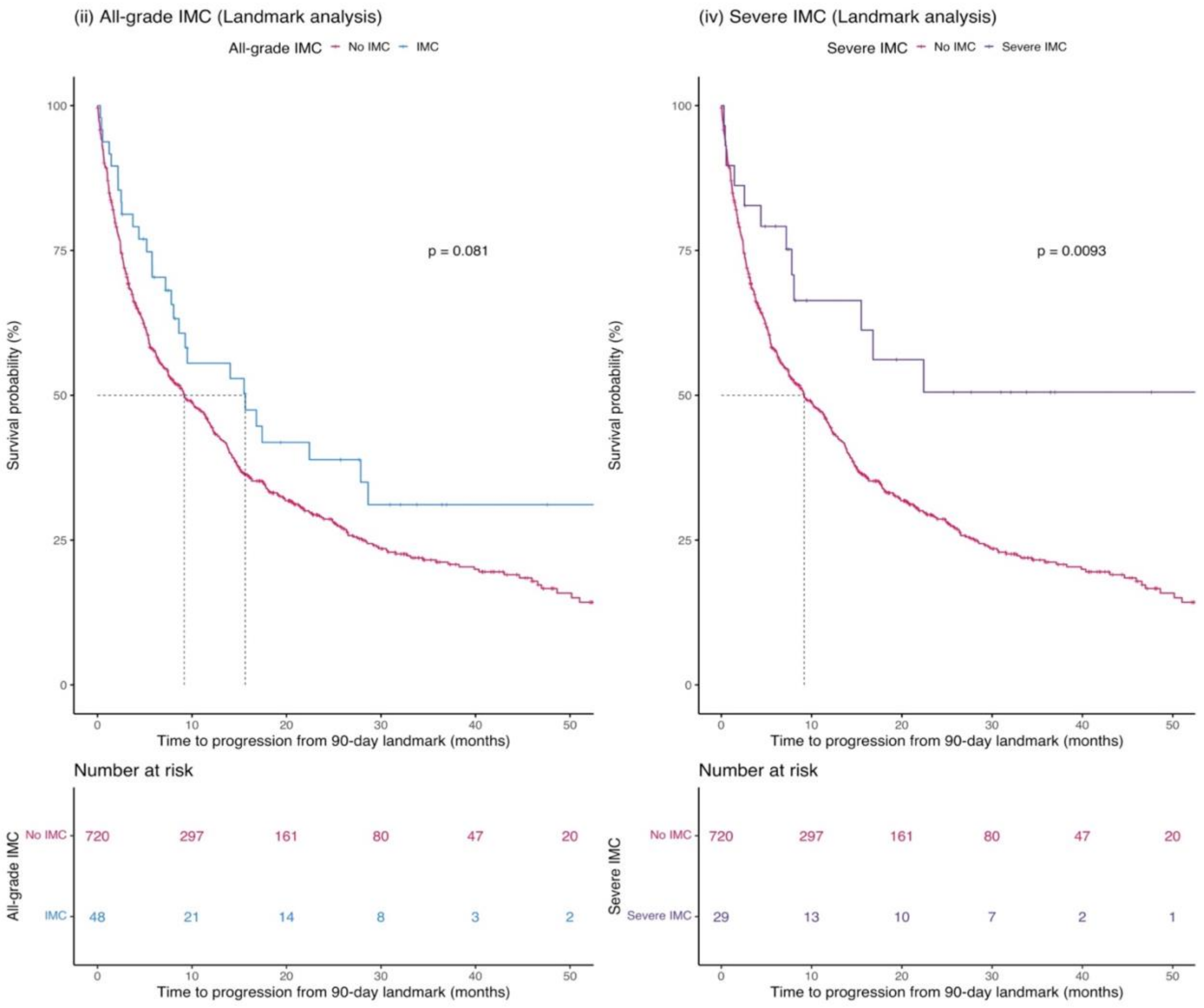
