## Supplementary Tables for "Polygenic risk score for ulcerative colitis predicts immune checkpoint inhibitor-mediated colitis"

**Supplementary Table 1: Characteristics of the BioVU replication cohort overall and by immune checkpoint inhibitor-mediated colitis (IMC)**

| Characteristics | Overall | All-grade IMC | Severe IMC |
|---|---|---|---|
| | (n=1147) | (n=104) | (n=83) |
| **Mean age at diagnosis (SD)** | 62.1 (13.1) | 61.6 (12.7) | 61.4 (12.9) |
| **Sex, n (%)** | | | |
| *Male* | 727 (63.4) | 62 (59.6) | 48 (57.8) |
| *Female* | 420 (36.6) | 42 (40.4) | 35 (42.2) |
| **Type of therapy, n (%)** | | | |
| *Anti PD-1/PD-L1 monotherapy* | 828 (72.7) | 34 (32.7) | 23 (27.7) |
| *Anti PD-1/PD-L1 + Anti CTLA4 therapy* | 45 (3.9) | 25 (24.0) | 22 (26.5) |
| *Anti CTLA4 monotherapy* | 274 (23.9) | 45 (43.3) | 38 (45.8) |

IMC: Immune checkpoint inhibitor-mediated colitis, SD: Standard deviation

**Supplementary Table 2: Previously published polygenic risk score (PRS) of ulcerative colitis as a predictor of time to development of all-grade immune checkpoint inhibitor-mediated colitis (IMC) in the entire GeRI cohort, using Cox proportional hazards models**

| | | *All-grade IMC* | | | |
|---|---|---|---|---|---|
| ***PRS[a]*** | ***PRS method*** | **HR per SD** | **95% CI** | ***P*** | ***Ref.*** |
| 179-SNP PRS | SNPnet | 1.24 | 0.94 - 1.64 | 0.13 | 1 |
| 809-SNP PRS | SNPnet | **1.33** | **1.01 – 1.74** | **0.04** | 1 |
| 1,505-SNP PRS | Penalized Regression | **1.37** | **1.02 – 1.85** | **0.04** | 2 |
| 566,637-SNP PRS | LDPred2 | **1.41** | **1.04 – 1.91** | **0.03** | 2 |

[a]Models are adjusted for age at diagnosis, sex, histology, type of therapy, recruiting site, and 5 principal components. IMC: Immune checkpoint inhibitor-mediated colitis, PRS: Polygenic risk score, HR: Hazard ratio, SD: Standard deviation, CI: Confidence interval, SNP: Single nucleotide polymorphism


1. Tanigawa Y, Qian J, Venkataraman G, et al. Significant sparse polygenic risk scores across 813 traits in UK Biobank. *PLOS Genetics*. 2022;18(3):e1010105. doi:10.1371/journal.pgen.1010105

2. Privé F, Aschard H, Carmi S, et al. Portability of 245 polygenic scores when derived from the UK Biobank and applied to 9 ancestry groups from the same cohort. *The American Journal of Human Genetics*. 2022;109(1):12-23. doi:10.1016/j.ajhg.2021.11.008
